# High correlation between binding IgG (anti-RBD/S) and neutralizing antibodies against SARS-CoV-2 six months after vaccination

**DOI:** 10.1101/2021.12.10.21267607

**Authors:** Raquel Guiomar, Ana João Santos, Aryse Martins Melo, Inês Costa, Rita Matos, Ana Paula Rodrigues, Irina Kislaya, Anabela Santos Silva, Carla Roque, Carla Silva, Joaquim Aguiar, Fátima Graça, Antônio Silva Graça, Ausenda Machado

## Abstract

Vaccination is considered the most important measure to control the COVID-19 pandemic. Extensive follow-up studies with distinct vaccines and populations are able to promote robust and reliable data to better understand the effectiveness of this pharmacologic strategy. In this sense, we present data regarding binding and neutralizing antibodies throughout time, from vaccinated and previously infected (PI) health care workers (HCW) in Portugal. We analyzed serum samples of 132 HCW, vaccinated and with previous SARS-CoV-2 infection. Samples were collected before vaccination (baseline, M1), at second dose vaccine uptake (M2), and 25-70 days (M3) and 150-210 days (M4) after the second dose for vaccinated individuals. The IgG (anti-RBD/S) antibody geometric mean titer found on vaccinated HCW at M2 (814.7 AU/ml; 95% CI 649.8-1021.5) were significantly higher than those found on PI HCW at recruitment (M1) (252.6 AU/ml; 95% CI 108.7 - 587.1), and the neutralizing antibodies (nAb) were similar between these groups, 93.2 UI/ml (95% CI 73.2-118.5) vs. 84.1 UI/ml (95% CI 40.4-155.9), respectively. We detected about 10-fold higher IgG (anti-RBD/S) antibodies titers in M3 when compared with M2, with a slightly but significant decrease in titers from 36 days after the second dose vaccine uptake. The increase of nAb titers were correlated with IgG (anti-RBD/S) antibodies titers, however, contrasting to IgG (anti-RBD/S) antibodies titers, we did not detect a decrease in nAb titer from 36 days after a second vaccine dose uptake. At M4, was observed a decrease of 8-fold in binding IgG (anti-RBD/S) and nAb. No significant differences in antibody titers were observed by sex, age or chronic diseases. Our results suggest that IgG (anti-RBD/S) antibodies titers and nAb titers could be correlated, but ongoing follow up of the cohort, is required to better understand this correlation, and the duration of the immune response.

## 1. Introduction

Vaccination is an important public health measure to control the Coronavirus Disease 2019 (COVID-19) caused by the Severe Acute Respiratory Syndrome Coronavirus 2 (SARS-CoV-2). Substantial effort has been taken worldwide to develop vaccines able to protect against COVID-19 [1]. Multiple mech-anisms were used in vaccines development, among them, the novel technology of mRNA-based vaccines [2], such as Comirnaty® and Spikevax® and the adenoviral vector vaccines as Vaxzevria®, and COVID-19 Vaccine Janssen® which are being used in the vaccination program of Portugal, to date [3]. In the actual pandemic context, given the record time from the research to the production and mass application of vaccines, the follow-up of vaccinated people is essential to obtain data regarding antibody response among different populations, under different epidemiological contexts [4]. In addition to the vaccine-effectiveness estimates, studies on immunogenicity are important to monitor vaccine performance.

The Spike protein (S) of SARS-CoV-2 is actually the target used in most vaccine development, since the receptor-binding domain (RBD), in subunit S1 of this protein, is considered the main target to binding and neutralizing antibodies [5]. Thus, it is expected that vaccinated people present anti-protein S antibodies, while people with previous COVID-19 may present anti-protein S and anti-nucleo-protein antibodies [6].

Previous studies demonstrated high seroconversion (up to 90%) in both vaccinated and infected people, although high heterogeneity among individuals was observed [7–9], however, the duration of the adaptive immune response is not well established to date. Some studies have demonstrated antibodies persistence in previously infected individuals for, at least, 12 months after symptoms onset [7], and at least 6 months after the complete vaccination scheme in vaccinated individuals [10].

The correlation between binding and neutralizing antibodies titers and protection is not clear in infection by SARS-CoV-2, and a cut off to predict protection is not available. A meta-analysis study based on 7 different COVID-19 vaccines had evidenced a correlation between binding and neutralizing antibodies with protection against symptomatic COVID-19, indicating the use of antibodies tests to correlate protection against disease, but no threshold was established [11].

In order to clarify questions regarding heterogeneity of immune response among individuals, duration of the adaptive immune response, correlation among biding and neutralizing antibodies and protection, the future need of vaccines boosters, among others, it is essential to carry on studies based on real-life observation of different populations and on different vaccines. This is particularly true for serological response and vaccine effectiveness measured at same time. The answers to these questions are an important key to delineate the next strategies to control the COVID-19 pandemic.

In Portugal, the vaccination program against COVID-19 started in December 27, 2020. As in the majority of European countries, the National Directorate of Health and the Ministry of Health defined a strategy that prioritized front-line Health Care Workers (HCW) for vaccination, since they are essential to maintain the health care units operational during the pandemic, but also because they are at higher risk of SARS-CoV-2 infection, and given the closed contact between these professionals and patients with comorbidities, with increased possibility of virus transmission to patients with high risk for severe disease [12]. The first vaccines to be available in Portugal were Comirnaty® followed by Spikevax® (both vaccines recommended to ≥12 years); Vaxzevria® (first recommended to individuals aged less than 65 years and then changed into individuals aged 60 or more years) and COVID-19 Vaccine Janssen® (recommended to women aged ≥50 years old or adults men) [3]. Following the national vaccination guidelines, during the study period those people with previous infection were vaccinated after 6 months from the laboratory diagnosis, with one dose of any vaccine [13].

The National Institute of Health Dr. Ricardo Jorge (INSA) has approximately 500 HCW in its staff, into which 81 were considered as front-line HCW to receive the vaccine against COVID-19 in the first phase of the national vaccination program.

Assuming the importance of specific SARS-CoV-2 antibodies as a proxy of the immune response to COVID-19 vaccines, HCW of INSA were followed-up for the first 6 months after the second vaccine dose uptake. In this study, we report findings regarding binding and neutralizing antibodies in vaccinated and previously infected HCW of INSA, in Portugal, from the baseline moment (recruitment or the first vaccine dose uptake) throughout, 6 months after the second vaccine dose uptake.

## 2. Materials and Methods

### 2.1. Design and participants

A prospective cohort study among INSA’s staff was implemented to examine SARS-CoV-2 vaccine effectiveness, including the serological component.

The INSA institutional vaccination campaign began on 12 January 2021. HCW (aged ≥18 years old) were invited to participate and were assigned into either the vaccinated cohort or the non-vaccinated cohort at the beginning of the follow-up period. Individuals were asked to participate via institutional email by the occupational medical service.

At the baseline, all participants filled-in a recruitment questionnaire were risk factors, symptoms and vaccination data were collected. In addition, all participants were followed on a weekly basis, by fill-in an online questionnaire on SARS-CoV-2 exposure and infection symptoms. Nasopharyngeal/oro-pharyngeal swab was collected for SARS-CoV-2 detection by RT-PCR tests when reported suspected signs and symptoms of COVID-19 on the weekly questionnaire or under the periodic testing screening at INSA.

### 2.2. Inclusion criteria

All staff of INSA that consent to participate of our cohort study of vaccine effectiveness against COVID-19 was included in this research project.

### 2.3. Exclusion criteria

The participants of our cohort study that did not received any dose of vaccine or that had not a previous infection were excluded of this analysis.

### 2.4. Definitions

We considered as vaccinated, individuals with more than 14 days after complete vaccination (receiving all doses recommended in the product characteristics); and as partially vaccinated, individuals with 14 days after receiving the first dose and until 14 days after receiving the second dose (in case of 2 doses). Additionally, as “previous infection” an individual who reported a positive RT-PCR test to SARS-CoV-2 or had a positive RT-PCR test to SARS-CoV-2 and/or had anti-SARS-CoV-2 antibodies in a serum sample taken in the recruitment at Moment 1(M1), before vaccine uptake.

### 2.5. Serological tests

Blood samples (3-5mL) were collected through venipuncture at baseline (M1) for all participants (including previously infected individuals), 30 days after first dose vaccine uptake (M2), 30 days after second dose (M3) and at 6 months follow up (M4). After a centrifugation at 2500rpm for 15min serum was obtained and conserved refrigerated (2 – 8ºC) for a maximum of 7 days before laboratorial analysis to detection of IgG antibodies and then, were stored frozen (−20ºC) until the neutralizing antibodies test was performed.

### 2.6. Determination of IgG antibodies

The determination of SARS-CoV-2 specific antibodies was done by a chemiluminescence enzyme immunoassay used for quantitative detection of Antireceptor-binding domain (RBD) from spike protein (S) antibodies (IgG anti-RBD/S) against SARS-CoV-2. Assays were performed in serum samples by the SARS-CoV-2 IgG II Quant assay (Abbott Diagnostics, IL, USA). Sera samples were considered positive when presented results >50 AU/mL. The tests were performed according to the manufacturer’s instructions.

The determination of IgG(anti-RBD/S) was performed at baseline, before first vaccine dose uptake (M1), before second vaccine dose uptake (M2), 25 to 70 days after second vaccine dose or completion of vaccination scheme (M3), and at 150 and 210 days after second vaccine dose or completion of vaccination scheme (M4).

### 2.7. Determination of neutralizing antibodies

The determination of neutralizing antibodies was performed at M2, M3 and M4 to vaccinated participants and at M1 to participants that had previous infection, using the commercial Enzyme-Linked Immunosorbent Assay (ELISA) kit TECO® SARS-CoV-2 Neutralization Antibody Assay (TECOmedical AG, Sissach, Switzerland), in the fully automated ELISA System DYNEX DS2®(Chantilly, VA, USA). The test has the principle of competitive binding, based on protein-protein interaction from the virus spike (S) protein (receptor binding domain - RBD) and the host cell receptor protein (angiotensin-converting enzyme 2 -ACE2). The test was performed according to the manufacturer’s instructions. The interpretation of the results was performed by the DYNEX DS2® system, and the results in IU/mL was obtained based on the optic density (OD) of each sample. The limit of interpretation of the equipment is in the interval between 5 IU/mL and 500 IU/mL. Those samples that presented a value >500 IU/mL to nAb were reanalyzed in dilutions of 5X or 10X, to obtain the most robust result possible to perform the statistical correlation among binding and neutralizing antibodies. The cutoff to determine the presence of neutralizing antibodies was stablished as >20 IU/mL.

### 2.8. Statistical analyses

Demographic, social and health characteristics of vaccinated (including partially vaccinated) individuals at baseline are described as relative frequencies for categorical and means and standard deviations for numerical variables.

For vaccinated, partially vaccinated and individuals with previous infection of COVID-19 data on the quantification of IgG (anti-RBD/S) and nAb antibody response activity was represented as an estimated geometric mean (GM) with 95% confidence interval (95% CI). The Wilcoxon’s rank sum test was applied to detect statistical differences for the IgG (anti-RBD/S) titers (GMT) at different moments. Between the 1st dose (20-30 days after vaccination, M2) and 2nd vaccine dose or completion of vaccination scheme (25-70 days after vaccination, M3), and between these two moments and 5 to 7 months after vaccination (150 to 210 days after 2nd dose or 1st dose in one uptake vaccine scheme, M4). Given the difference of elapsed time since completion of vaccination scheme and blood sample collection at the M3, we also tested the differenced in GMT for two moments after vaccination scheme completion (25-35 days vs. 36-70 days). Spearman’s coefficient and p-value were calculated to evaluate the correlation between IgG (anti-RBD/S) and neutralizing response activity (nAb). A linear regression was performed on log-transformed IgG (anti-RBD/S) titers at the M3 (25 to 70 days after 2nd vaccine dose) and at the M4 (150 to 210 days after 2nd vaccine dose) to determine the association to sex, age groups (20-39, 40-70 years) or chronic disease (at least one chronic disease/no disease) for vaccinated individuals. Statistical significance was defined as p < 0.05.

All statistical analyses were performed using Stata software, version 15 (StataCorp.2017. Stata Statistical Software).

### 2.9. Ethical considerations

The study complied with legal and ethical requirements. The study protocol was approved by the National Institute of Health Doctor Ricardo Jorge Health Ethics Committee. All participants provided written informed consent for collection of data regarding demographic, social, and health information, blood samples and nasopharyngeal/oropharyngeal swabs.

## 3. Results

Out of 212 workers that integrated the INSA cohort at 15 June 2021, 132 were included in this study: 114 from the vaccinated and partially vaccinated group and 18 individuals with previous infection: with a RT-PCR positive test (n=14) or with a positive serological analysis with the detection of antibodies against SARS-CoV-2 in serum sample (n=4). These 4 individuals were therefore unaware of having been expose to the SARS-CoV-2.

From the vaccinated group, 84 were vaccinated against SARS-CoV-2, and 30 were partially vaccinated (Table 1). Vaccinated participants had mostly taken the Comirnaty® vaccine (96.4%), the others with COVID-19 Vaccine Janssen® (2.4%) and the Vaxzevria® (1.2%). Of the partially vaccinated individuals, 66.7% had taken the Comirnaty®, 20% the Vaxzevria® and 13.3% the Spikevax. In both groups, the majority were women (81% and 90%, respectively), and the mean of ages was lower for the vaccinated group (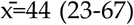 versus 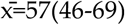). Within the vaccinated group, over 68% reported to work in laboratory, whereas over 81% of the partially vaccinated group reported to work in services that did not required interaction with the public. Regarding smoking habits, 11% of the fully vaccinated individuals and 23% of the partially vaccinated reported to be smokers. Little less than half of vaccinated (48.7%) and 68.4% of the partially vaccinated participants reported at least one chronic disease. In both groups, over 70% of participants reported to have uptake the influenza vaccine in the previous season.

**Table 1.** Vaccinated and partially vaccinated participants’ sociodemographic, work, and health characteristics.

|  |  | Vaccinated |  | Partially vaccinated<br>(1 dose)* |  |
| --- | --- | --- | --- | --- | --- |
|  |  | n | % | n | % |
| **Total** |  | 84 |  | 30 |  |
| **Sex** |  |  |  |  |  |
|  | Female | 68 | 81.0 | 27 | 90.0 |
|  | Male | 16 | 19.0 | 3 | 10.0 |
| **Age, mean [range]** |  | (x̄=44[23-67]) |  | (x̄=57[46-69]) |  |
| **Age groups (n)** |  | (83) |  | (30) |  |
|  | 20 to 49 years | 44 | 53.0 | 5 | 16.7 |
|  | 50 to 70 years | 39 | 47.0 | 25 | 83.3 |
| **Work functions** |  | (79) |  | (26) |  |
|  | Without public contact | 13 | 16.5 | 21 | 80.8 |
|  | Public contact | 12 | 15.2 | 4 | 15.4 |
|  | Laboratory | 54 | 68.4 | 1 | 3.8 |
| **Smoking** |  | (82) |  | (30) |  |
|  | Never smoked | 54 | 65.9 | 16 | 53.3 |
|  | Smoker | 9 | 11.0 | 7 | 23.3 |
|  | Former smoker | 19 | 23.2 | 7 | 23.3 |
| **Chronic disease (n)** |  | (70) |  | (19) |  |
|  | No disease | 38 | 54.3 | 6 | 31.6 |
|  | 1+ disease | 32 | 45.7 | 13 | 68.4 |
| **Flu vaccine season**<br>**2020/2021 (n)** |  | (81) |  | (28) |  |
|  | No | 24 | 29.3 | 5 | 17.9 |
|  | Yes | 58 | 70.7 | 23 | 82.1 |

At M1, the IgG (anti-RBD/S) titers were higher for individuals with previous SARS-CoV-2 infection (**Erro! A origem da referência não foi encontrada**.). Information about the date of the positive PCR test was only available for 11 of the 18 individuals with previous infection. Individuals that have had previous infection in the last 90 days presented a higher GM (GM=627.5AU/mL; CI: 168.3-2338.8) than those individuals that had been infected prior to 90 days (GM=408.5 AU/mL; CI: 184.2-905.9).

**Figure 1.**
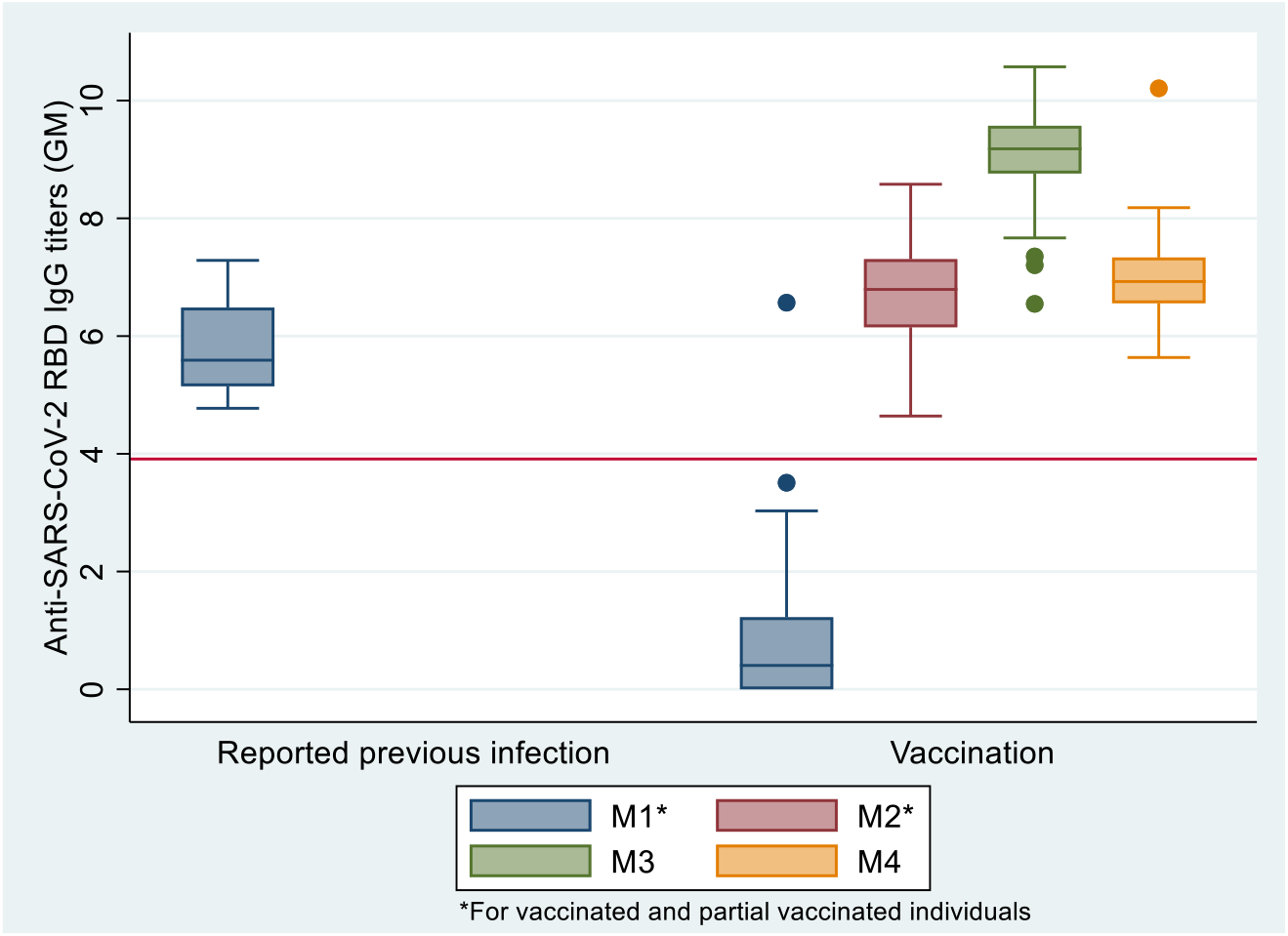
Concentration of IgG anti-SARS-CoV-2 spike receptor-binding domain (RBD) titers reported in the box–whisker plots for individuals with previous infection and vaccinated individuals without previous infection for the four different moments of observation.

Table 2 displays the GM for IgG (anti-RBD/S) titers for different groups and moments. The concentration of IgG (anti-RBD/S) was significantly higher (GM= 814.7 AU/mL; CI: 649.8-1021.5) for the vaccinated individuals at M2, when compared to the values observed at M1 for the individuals previously infected (GM= 252.6 AU/mL; CI:108.7-587.1) (p<0.001).

**Table 2.** Geometric Mean (GM) of SARS-CoV-2 IgG (anti-RBD/S) concentration titers at four different moments for the full-vaccinated, partially vaccinated participants and at moment of recruitment for individuals with previous infection.

| Groups | M1<br>[CI95%] (n) | M2<br>(Vaccinated<br>and partial)<br>[CI95%] (n) | M3 (Vaccinated) |  |  | M4<br>(Vaccinated)<br>[CI95%] (n) |
| --- | --- | --- | --- | --- | --- | --- |
|  |  |  | Total<br>[CI95%] (n) | 25-35 days<br>[CI95%] (n) | 36-70 days<br>[CI95%] (n) | 150-210 days<br>[CI95%] (n) |
| Vaccinated<br>& Partially<br>vaccinated* | 2.5 [1.9-3.4]<br>(n=96) | 814.7 [649.8-1021.5] (n=63) | 8803.8[7529.5<br>10293.7] (n=81) | 10409.0 [8507.9<br>12734.7] (n=49) | 6812.2 [5400.7<br>8592.6] (n=32) | 1070.4 [922.5-1241.9] (n=72) |
| **Previous infection (n=17)** | **252.6 [108.7**<br>**587.1]** | – | – | – | – | – |

For vaccinated participants, antibodies titers were significantly higher after the second vaccine dose, with an increase of about 10-fold from M2 to M3 in the IgG (anti-RBD/S) antibodies titers (GM=8803.8AU/mL, 95 % CI: 7529.5-10293.7) (*p*<0.0001). For the M3, serum samples were drawn at several distinct times, comprising a broad range of days between sample collection and second dose uptake. Vaccinated individuals whose blood samples were drawn between 25 and 35 days after the second dose presented a higher GM of IgG (anti-RBD/S) concentration (GM=10773.0AU/mL, CI:8910.2-13025.3), compared to those whose sample collection took place after the 35 until 70 days (6912.1AU/mL, CI:5447.3-8770.8) (p<0.05). Even though the concentration of IgG (anti-RBD/S) titers decreased at M4 (GM= 1070.4 AU/mL; CI: 922.5-1241.9) and was significantly lower when compared to the values observed at M3 (p<0.001). Antibody titers at M4 were still significantly higher than observed for the vaccinated individuals at M2, after the 1^st^ vaccine dose (p<0.05).

Linear regression was performed on log-transformed IgG (anti-RBD/S) titers at the M3 and M4 to determine the association to sex, age groups or chronic disease for vaccinated individuals (Table 3). No statistical differences (p>0.05) in the GM of IgG (anti-RBD/S) concentration were observed between men or women, between individuals in the two age groups or between individuals without or with at least one chronic disease at each of the observed moments.

**Table 3.** Regression on log-transformed SARS-CoV-2 IgG (anti-RBD/S) titers at the M3 (25-70 days after 2^nd^ dose) and M4(150-210 days after 2^nd^ dose) for full-vaccinated individuals.

| Variable | M3 |  |  | M4 |  |  |
| --- | --- | --- | --- | --- | --- | --- |
|  | Odds Ratio | 95% CI | P | Odds Ratio | 95% CI | P |
| **Sex** | (n=87) |  |  | (n=72) |  |  |
| Female | * |  |  | * |  |  |
| Male | 0.83 | 0.56-1.2 | 0.358 | 0.79 | 0.51-1.2 | 0.298 |
| **Age** | (n=86) |  |  | (n=72) |  |  |
| 20-49 | * |  |  | * |  |  |
| 50-70 years | 1.08 | 0.79-1.48 | 0.626 | 0.86 | 0.60-1.2 | 0.415 |
| **Chronic disease** | (n=71) |  |  | (n=61) |  |  |
| No disease | * |  |  | * |  |  |
| 1+ disease | 1.19 | 0.84-1.69 | 0.310 | 0.95 | 0.70-1.3 | 0.776 |

\*Constant

To explore the humoral immune response generated after SARS-CoV-2 infection, the neutralizing antibody response (nAb) was evaluated for individuals with previous infection at recruitment (M1), and for vaccinated individuals at M2, M3 and M4 (Figure 2).

**Figure 2.**
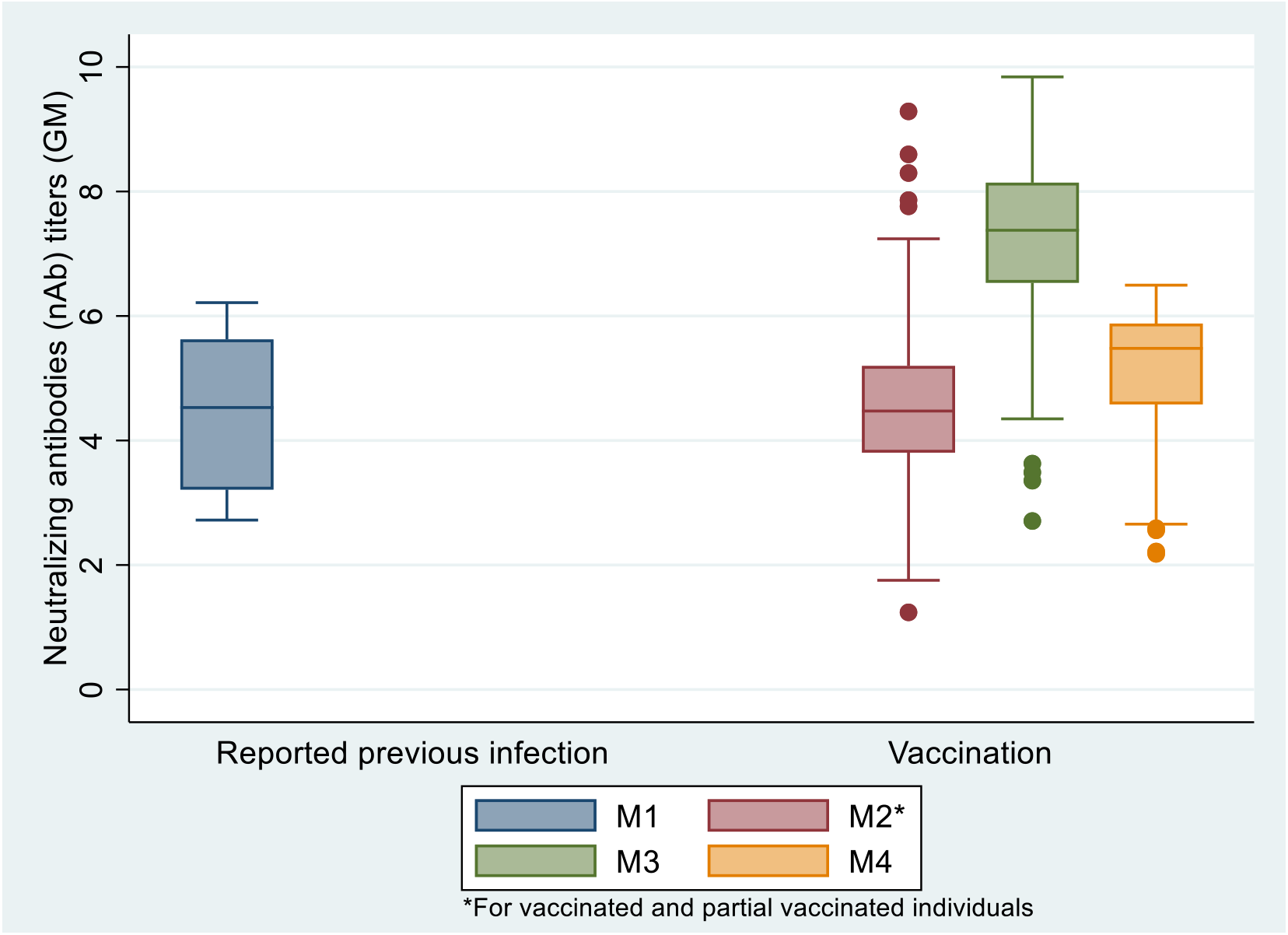
Concentration of neutralizing antibodies (nAb) titers reported in the box–whisker plots for individuals with previous infection and vaccinated individuals without previous infection for the four different moments of observation.

Table 4 presents the geometric mean of nAb activity in serum samples of participants with previous infection at M1, and for vaccinated and partially vaccinated individuals at M2 and M3. The GM of nAb in previously infected individuals at M1 (GM=84.1 mL; CI: 40.4-155.9) was not different from that observed for vaccinated individuals at M2 (GM=93.2IU/mL; CI: 73.2-118.5) (p=0.645). On the other hand, when compared with M2, a significant increase of nAb titer was observed in the M3 for vaccinated individuals (GM=1267.3 IU/mL; CI: 1060.6-1514.4) (p<0.001). Statistical significant differences were also observed in the nAb concentrations between 25 to 35 days (GM=1551.9 IU/mL; CI: 1261.4-1909.5) and 36 and 70 days (GM=935.2 IU/mL; CI: 691.6-1264.7) after the second vaccine dose or completion of the vaccination scheme (p<0.05). Even though a decrease of nAb concentrations titters in the last moment of observation (M4, GM=165.8 IU/mL; CI: 128.4-214.2), the titers were still significantly higher than those observed after the first vaccine dose (p<0.05).

**Table 4.** Geometric Mean (GM) of neutralizing antibodies (nAb) titers at four different moments for the full-vaccinated, partially vaccinated participants and at moment of recruitment for individuals with previous infection

| Groups | M1<br>[CI95%] (n) | M2<br>(Vaccinated<br>and partial)<br>[CI95%] (n) | M3 (Vaccinated) |  |  | M4<br>(Vaccinated) |
| --- | --- | --- | --- | --- | --- | --- |
|  |  |  | Total<br>[CI95%] (n) | 25-35 days<br>[CI95%] (n) | 36-70 days<br>[CI95%] (n) | 150-210 days<br>[CI95%] (n) |
| **Vaccinated**<br>**& Partially**<br>**vaccinated\*** | – | 93.2[73.2-118.5] (n=84) | 1267.3 [1060.6<br>1514.4] (n=80) | 1551.9 [1261.4-1909.5] (n=48) | 935.2 [691.6<br>1264.7] (n=32) | 165.8[128.4-214.2] (n=68) |
| **Previous**<br>**infection**<br>**(n=14)** | 84.1 [40.4-155.9] | – | – | – | – | – |

High statistically significant correlations were observed between IgG (anti-RBD/S) and nAb titers, at all observational moments. After the first vaccine dose (M2) IgG (anti-RBD/S) titers correlated highly with nAb titers (Spearman’s ρ = 0.79, p<0.001). Similar higher correlations were observed at M3 (Spearman’s ρ = 0.86, p<0.001), either 25 to 35 days (Spearman’s ρ = 0. 81, p<0.001) or 36 to 70 days after completion of vaccination scheme (Spearman’s ρ = 0. 87, p<0.001). Finally, at M4, a significant correlation, was observed between IgG (anti-RBD/S) and nAb titers (Spearman’s ρ = 0.70, p<0.001).

## 4. Discussion

In this work, we accessed the preliminary data regarding binding and neutralizing antibodies in HCW from National Public Health Institute, Portugal, from the baseline, before first vaccine dose uptake, until 150-210 days (5-7 months) after the second vaccine dose. We demonstrated that the first vaccine dose elicited an immunological response although a second dose was essential to promote a boost in binding and neutralizing antibodies SARS-CoV-2. IgG (anti-RBD/S) were highly correlated with neutralizing antibodies, being higher in the first 70 days (10 weeks), keeping however a good correlation after completion of the vaccination scheme.

The IgG (anti-RBD/S) titer was significantly higher in vaccinated individuals, after one or two vaccine doses, compared with baseline values of individuals that had developed an immunity response after a previous SARS-CoV-2 infection. The GM of IgG (anti-RBD/S), after one vaccine dose, from vaccinated individuals was significantly higher (p<0.001) than GM found at recruitment for individuals that had a previous infection. This data is in accordance with the previously reported from a cohort of HCW from an academic medical center in Southern California [14] and with the Portuguese national serological survey performed in February-March 2020 [15]. However, we have to take in consideration that the elapsed time between infection and the recruitment to this study had a great variation among individuals. Due to the lower sample size between these groups, it is not possible to perform a robust comparative analysis in the group of previous SARS-CoV-2 infection.

The marked increase of the IgG (anti-RBD/S) after the second dose, found in this study was compatible with the observed in previous studies [6,8,10,14], and in trials studies[16]. It is important to highlight that after the second dose we found a significant decrease for IgG (anti-RBD/S) between individuals that had a serum analysis from 25 to 35 days and those that had a serum analysis from 36 to 70 days, after the second dose. This decrease was expected given that after the second dose there is a high stimulation of immune system, with high production of antibodies, however it is not expected that the IgG titers be kept at the maximum for a long time, and though a decreasing to basal titers of memory is generally observed [17,18]. This decrease was also found in other studies [10,19]. In this sense, is important to keep the follow up of the cohort studies in order to clear for how long the IgG titers remains and to try to correlate this data with nAb, and to establish a cut off that could predict protection against SARS-CoV-2 infection. Other studies in health care workers showed that the efficient immunological response to COVID-19 vaccines is associated with a reduction of new COVID-19 cases among those who received two doses of the vaccine, even when a surge of the B.1.1.7 variant was noted in up to 80% of cases. The effective vaccination among health care workers provides a safe environment, even in the presence of a high rate of SARS-CoV-2 infection in the community. Although there is a good effectiveness for COVID-19 vaccines, is recognized a decrease in vaccine effectiveness with time [20]. A study from England, found high levels of vaccine effectiveness against symptomatic disease after two doses, even when a new variant Delta was circulating [21]. New variants can also pose a challenge to COVID-19 vaccine effectiveness that expresses the perfusion stabilized full spike glycoprotein (S) of the original SARS-CoV-2 isolate Wuhan-Hu-1, but recent studies have already highlighted that variants of concern [22], Alpha (B.1.1.7 variant), Beta variant first identified in South Africa (B.1.351 lineage), and Gamma variant first identified in Brazil (P.1 lineage) remained susceptible to Comirnaty® vaccine elicited serum neutralization, although at a reduced level for the B.1.351 variant [23,24]. For the Delta variant, predominant in Portugal since mid-June 2021 [25], studies on cross-reactivity of monoclonal antibodies to pre-existing SARS-CoV-2 strains, showed that vaccination of previously infected individuals is likely to be protective against a large array of circulating viral strains, including the Delta variant. In the same way, two-dose regimen generated high sera-neutralization levels against the Alpha, Beta and Delta variants in individuals sampled at week 8 to week 16 after vaccination [26], being the levels of neutralizing antibodies highly predictive of immune protection from symptomatic SARS-CoV-2 infection [27]. A study at national level estimated high mRNA VE for the prevention of COVID-19-related hospitalizations and deaths (in ≥ 65 years, full vaccinated) with any evidence of VE reduction in the 3 months after the second dose uptake, during the period of Delta variant circulation [28], consistent with the high titers for binding and neutralizing antibodies detected after complete vaccination.

In opposite to other studies such as one performed in UK, where people older than 50 years old presented a weaker serological response in relation to those younger than 50 years old [29], in our study we did not identify any difference between age, sex, or the presence of one or more chronic diseases. This fact can be due to the limited sample size of our study, which do not allow a more robust analysis.

Our study has some limitations, the cohort includes only active healthy workers, without severe comorbidities, and all participants are under 70 years old, with over representation of the female population. The occupational risk is reduced, although the majority of the participants manipulate SARS-CoV-2 positive samples, strict guidelines to use individual protective equipment limit the occupational risk exposure being reduced compared to medical personnel with close contact with patients. The heterogeneity in the reference units to quantify the detected antibodies posed a difficulty to compare our data with other studies. Few studies used the same units that we used in this study. This point was already highlighted by Earle et al., [11] that suggested the use of the WHO International Standard (NIBSC 20/136) to express neutralizing antibodies titers in IU/mL and binding antibodies titers in BAU/mL to be possible to compare data among different studies using serology assays, in the context of COVID-19 pandemic. Given the importance to compare different vaccines among different populations to help to design new strategies to battle the pandemic, this point is crucial to a better comprehension of data obtained around the world. We didn’t explore the antibody titers for variants of concern, and a decrease of antibody titers could be expected. The cellular immunity and other immunological mechanisms weren’t explored during the study.

In our study, we found a high correlation between biding and neutralizing antibodies after the first dose and similar results have been previously reported by other authors [30,31]. It was reported that nAb remain relatively stable for several months after infection, but there is a lack of information on for how long time it persists [31]. The slower decrease in nAb after the second vaccine dose uptake, could support a robust and long persistence of nAB after full vaccination [20,31,32]. A recent study reported a good correlation between binding and neutralizing antibodies levels and protection against symptomatic infection [33]. Although we observed a decrease in nAb titers levels about 6 months after vaccination, it was high far away from the minimum level reported by Feng et al. associated with 80%VE against symptomatic infection, with majority Alpha variant (26 IU/mL for pseudovirus neutralization). In our study, we were not able to predict the loss of protection due to the decrease of nAb at 5-7 months after full vaccination neither to confirm that the nAb levels found at this moment are considered a robust immune response. In this sense, we recommend caution in the interpretation of the decrease of the levels of biding and neutralizing antibodies, especially in face to the new variants challenge.

The preliminary data obtained from our cohort study demonstrate the importance to keep the follow up of the individuals in order to better understand the behavior of immune response to the COVID-19 vaccines, and to try to establish a threshold that could predicts protection against the SARS-CoV-2 infection. Serological studies precluding the waning of antibody levels with time must be integrated in the vaccine effectiveness studies to better clear questions about the duration of immune protection and vaccine effectiveness. The data on waning immunity and vaccine effectiveness constitute important facts for health decision makers, to implement measures to reduced severe disease, mortality and transmission that could comprises non-pharmaceutical measures and/or additional booster vaccine doses.

## Data Availability

All data produced in the present study are available upon reasonable request to the authors

## Author Contributions

Conceptualization, R.G. and A.M..; methodology, A.M.M.; I.C.; R.M.; A.S.S.; C.R.; C.S.; J.A.; F.G.; A.S.G.; software, A.J.C.; I.K.; writing—original draft preparation, R.G; A.J.C.; A.M.M.; writing—review and editing, R.G.; A.J.C.; A.M.M.; I.C.; R.M.; A.P.R.; I.K.; A.S.S.; C.R:; C.S.; J.A.; F.G.; A.S.G.; A.M.. All authors have read and agreed to the published version of the manuscript.”

## Institutional Review Board Statement

The study was conducted according to the guidelines of the Declaration of Helsinki, and approved by the Ethics Committee of National Institute of Health Doutor Ricardo Jorge.

## Informed Consent Statement

Informed consent was obtained from all subjects involved in the study.

## Acknowledgments

We would like to acknowledge all the participants from the staff of the National Institute of Health for the availability to participate in the present study and to donate blood samples for the evaluation of the humoral immunity response after vaccination.

We acknowledge the colleagues that contributed to the support of the present study: Paula Braz from the Department of Epidemiology of INSA; Camila Henriques, Carla Manita, Fátima Martins, João Santos, Jorge Machado and Sofia Soeiro from the Infectious Diseases Department, INSA. We acknowledge the colleagues from Management and Laboratory Support Unit.

